# Protocol for a systematic review of qualitative and quantitative effects of cardiovascular disease risk communication using ‘heart age’ concepts

**DOI:** 10.1101/2020.05.03.20089938

**Authors:** Carissa Bonner, Carys Batcup, Samuel Cornell, Michael Anthony Fajardo, Jenny Doust, Kevin McGeechan, Lyndal Trevena

## Abstract

**Introduction:** The concept of ‘heart age’ is increasingly used for health promotion and alongside clinical guidelines for cardiovascular disease (CVD) prevention. These tools have been used by millions of consumers around the world, and many health organisations promote them as a way of encouraging lifestyle change. However, heart age tools vary widely in terms of their underlying risk models and display formats, the effectiveness of these tools compared to other CVD risk communication formats remains unclear, and doctors have raised concerns over their use to expand testing of healthy low risk adults.

**Methods and analysis:** We aim to systematically review both qualitative and quantitative evidence of the effects of heart age when presented to patients or consumers for the purpose of CVD risk communication. Four electronic databases will be search until April 2020 and reference lists from similar review articles will be searched. Studies will be considered eligible if they meet the following criteria: (1) published from the inception of the database to April 2020, in peer-reviewed journals, (2) used an adult population (over 18 years of age) or, if not explicit regarding age, are clear that participants were not children, (3) present the concept of ‘heart age’ to patients or consumers for the purpose of CVD risk communication, (4) report qualitative themes or quantitative outcomes relating to psychological and/or behavioural responses to heart age. Two reviewers will perform study selection, data extraction and quality assessment. Reporting of the review will be informed by Preferred Reporting Items for Systematic Review and Meta-Analysis guidance.

**Ethics and dissemination:** Ethical approval is not required as it is a protocol for a systematic review. Findings will be disseminated through peer-reviewed publications and conference presentations.

## Background

The concept of ‘heart age’ is increasingly used for health promotion and alongside clinical guidelines for cardiovascular disease (CVD) prevention.^1^ These tools have been used by millions of consumers around the world, and many health organisations promote them as a way of encouraging lifestyle change.^2–3^ However, heart age tools vary widely in terms of their underlying risk models and display formats, the effectiveness of these tools compared to other CVD risk communication formats remains unclear, and doctors have raised concerns over their use to expand testing of healthy low risk adults.^1,4^ A review of CVD risk communication in 2011 identified heart age as a potentially useful concept that needed more research.^5^ A 2016 review of vascular age concepts in clinical applications found limited trials testing the effects of communicating this concept,^6^ but a recent rapid review in 2020 highlighted the increasing number of studies on age-based risk formats, suggesting there may now be more evidence available.^7^

### Rationale for the systematic review

Earlier reviews relating to the heart age concept have not made a distinction between the comparison groups involved in trials (standard care or absolute risk), have not clearly identified the behaviour change techniques included within the heart age intervention, and/or have not included qualitative studies that can provide additional insights into why these tools are so widely used in spite of limited evidence for their effectiveness. We will systematically review both qualitative and quantitative evidence of the effects of heart age when presented to patients or consumers for the purpose of CVD risk communication.

## Aims and Objectives

### Review aim

The aim of the systematic review is to identify and review investigations which sought to determine the effectiveness of communicating ‘heart age’ to patients to change their lifestyle behaviours or which identified the feasibility of using ‘heart age’ to communicate risk of a cardiac event.

### Objectives

The primary objectives of the systematic review will be to review the qualitative and quantitative literature on the effects of communicating the ‘heart age’ concept to patients and consumers. This will be achieved by:

1. Describing the positive and negative effects of communicating heart age: using qualitative and quantitative research.
2. Exploring the reasons why heart age is psychologically impactful: using qualitative research.
3. Comparing the effect of heart age to other risk communication strategies: using quantitative RCTs.
4. Investigating what behaviour change techniques are included in the heart age interventions: using qualitative and quantitative research where full intervention content is accessible.

## Methods and Analysis

This protocol has been developed following the Preferred Reporting Items for Systematic Review and Meta-Analysis Protocols (PRISMA-P) guidelines. Reporting of the systematic review will be informed by Preferred Reporting Items for Systematic Review and Meta-Analysis guidance. Any relevant amendments made to the protocol will be documented and published alongside the results of the systematic review.

### Inclusion and exclusion criteria

#### Types of studies

Studies will be considered eligible if they meet the following criteria:

1. published from the inception of the database to April 2020, in peer-reviewed journals,
2. used an adult population (over 18 years of age) or, if not explicit regarding age, are clear that participants were not children,
3. present the concept of ‘heart age’ to patients or consumers for the purpose of CVD risk communication,
4. report qualitative themes or quantitative outcomes relating to psychological and/or behavioural responses to heart age.

Studies that are not peer reviewed journal articles such as; conference proceedings, dissertations or government reports will be excluded.

The study must include the heart age concept being communicated to patients or consumers. Therefore, protocol papers, opinion papers, reviews, online user descriptions and heart age algorithm development or validation will be excluded, and where the algorithm has been applied to population data but not presented to that population.

### Types of participants

All studies will involve an adult population. Studies using participants <18 years of age will be excluded, as will animal studies.

### Interventions

Any study that presents the concept of heart age to patients or consumers for the purpose of CVD communication, either alone or alongside other risk information. Interventions may be delivered in settings such as general practices, hospitals, health clinics or community centres, or online. Studies that do not report on heart age as a communication intervention will be excluded.

### Comparators

Investigations may include an intervention group with or without a control or comparator group, such as usual care in clinical practice or absolute CVD risk assessment alone.

### Information Sources

#### Electronic searches

We will search the following databases: The Cochrane Central Register of Controlled Trials (via OvidSP), Medline (via OvidSP), EMBASE (via OvidSP) and PsychINFO (via OvidSP) up to April 2020. The search terms are based on the earlier vascular age review in 2016 with additional free text terms based on known relevant papers. The full list of terms is based on a previous review and includes: vascular/vessel/arterial/heart/cardiovascular/coronary/risk + age/ages/ageing/aging; or heart forecast; and limited to human studies.

#### Searching other relevant resources

We will search the citations and references of final included studies and two previous reviews of vascular age models and more general age-related risk concepts. We will also include any papers mentioned on publicly accessible heart age websites.

### Study records

#### Data management

We will download references identified in searches (electronic databases and additional searches) into Excel. Duplicates will then be removed, and a copy of this spreadsheet will be made to enable two reviewers to screen the titles and abstracts of each study included.

#### Selection process

The screening process will be undertaken by two review authors individually. Each reviewer will independently assess a study’s suitability to be included in the review by way of marking as such against each study on an excel spreadsheet, which will contain the title and abstract. Studies that do not meet the inclusion criteria will be excluded at this stage. We will obtain the full text of the remaining papers and will then assess these remaining papers against the full inclusion terms for the review to determine their eligibility for inclusion. Non-English language papers will be translated into English using Google Translate and verified for inclusion/exclusion by speakers of the relevant language. The review authors will resolve disagreements through a consensus-based decision-making process, or when necessary, discussion with a third review author.

#### Data extraction process

Two review authors will use a pre-defined data extraction form to collect data from the included studies. Reviewers will pilot the data extraction form with a sample of included papers and amendments will be made if necessary. Reviewers will contact study authors if additional information is required. An Excel database will be used to extract quantitative and qualitative data from the included studies as follows:

***Quantitative data to be extracted will include:*** year, country, study design, study population (age, education, socio-economic status, health literacy, race/culture/ethnicity), number of participants, intervention format (online, paper), comparison groups (standard care, absolute risk alone), clinical measures (blood pressure, cholesterol, weight, body mass index, waist circumference, prescribed medications) behavioural measures (medication adherence, lifestyle intentions or self-report), psychological measures (risk perceptions, emotional responses, credibility, recall), summary of significant effects of heart age communication.

***Qualitative data to be extracted will include:*** behavioural themes (e.g. lifestyle change), psychological themes (e.g. emotional reaction), stated benefits of heart age (e.g. motivates people to take action), stated problems with heart age (e.g. reduces credibility).

***Intervention content data will include:*** additional risk communication formats (e.g. absolute risk, risk level, graphs), underlying model (e.g. 5 year or 10 year risk CVD risk model), behaviour change techniques (e.g. email prompts, action plans) based on Michie et al.’s taxonomy.^8^ This will be done for interventions where the full content is accessible, either within the publication, online or through contacting the authors.

#### Assessment of Risk of Bias in included studies

Studies will be critically appraised independently by two review authors using the relevant Johanna Briggs Institute tools for the study design (https://joannabriggs.org/ebp/critical_appraisal_tools). Disagreement will be resolved by discussion and the involvement of a third reviewer if necessary. Authors will be contacted if further information is required to make an assessment. Sensitivity analyses will be performed on included studies with a high risk of bias. Funnel plots will be generated to assess publication bias only if at least ten studies with comparable quantitative outcomes can be included in a meta-analysis.

## Outcomes and Prioritisation

To meet our research aims and objectives, the outcomes of interest for this study include the following:

1. Lifestyle risk factors for CVD (diet, exercise, smoking)
2. Clinical risk factors for CVD (blood pressure, cholesterol, weight-related measures)
3. Risk perceptions (perceived risk accuracy, severity, susceptibility, perceived control)
4. Emotional response (positive e.g. happy/motivated, negative e.g. sad/anxious)
5. Recall (exact heart age result, general result being older/younger than current age)
6. Perceived credibility (how believable the heart age result is)
7. Information seeking behaviour (accessing further information, seeing a doctor)

## Conclusion

This review will update previous related reviews with a more comprehensive list of search terms, the inclusion of both qualitative and quantitative studies, and more detailed analyses of the nature of heart age interventions in terms of the behaviour change techniques included alongside the heart age risk result itself.

## Data Availability

All data included in the systematic review will be publicly accessible online through journal databases.

